# Locus-specific stratification and prioritization unveil high risk genes underlying hyperuricemia

**DOI:** 10.1101/2024.03.06.24303846

**Authors:** Jing Zhang, Yue Guo, Luyu Gong, Limei Xia, Qiaoqiao Liu, Kangchun Wang, Qi Wang, Zhaojun Liu, Zhaohui Qin, Shaolin Shi, Jingping Yang

**Affiliations:** State Key Laboratory of Pharmaceutical Biotechnology, Medical School, Nanjing University, Nanjing, Jiangsu, 210093, China; Jiangsu Key Laboratory of Molecular Medicine, Medical School, Nanjing University, Nanjing, Jiangsu, 210093, China; Department of Biostatistics and Bioinformatics, Rollins School of Public Health, Emory University, Atlanta, GA, 30322, USA; National Clinical Research Center for Kidney Diseases, Jinling Hospital, Affiliated Hospital of Medical School, Nanjing University, Nanjing, Jiangsu, 210093, China

## Abstract

The development of alternative medications for urate-lowering therapies is imperative for patients that are intolerant to current treatments. Despite GWAS have identified hundreds of loci associated with serum urate levels, the mechanistic understanding and discovery of drug targets remain difficult. This difficulty arises from the multiple-independent-associations challenge in the genomic studies of complex diseases as hyperuricemia. Here, we introduced a locus-specific stratification (LSS) and gene regulatory prioritization score (GRPS) approach to address the multiple-independent-associations challenge. By integrating with kidney single-cell chromatin accessibility and gene expression, LSS identified functional SNPs, regulatory elements, and genes for 118 loci. The interpretability was increased by 1.4 to 5.2 fold. GRPS prioritized genes and nominated under-explored drug target with high confidence, which was validated using CRISPR activation and phenotypic assays. Our findings not only identified top causal genes but also proposed the regulatory mechanisms for pathogenic genes, expanding our knowledge of the genetic contribution in complex diseases as hyperuricemia.

**One-sentence summary:** A novel approach to comprehensively explore genetic contribution and nominate reliable causal genes for complex diseases as hyperuricemia.

## Introduction

Hyperuricemia, characterized by elevated serum urate levels, is a complex disease affecting various complications (*1–3*). Although the dominant narrative about hyperuricemia has focused on dietary, recent research suggests that diet plays only a limited role in regulation of serum urate in the healthy population (*4*). Instead, it has been estimated that genetic contribution for hyperuricemia ranges from 40% to 73% (*5*). There are currently urate-lowering therapies developed based on genetic targets, such as allopurinol, which inhibits xanthine oxidase and the production of urate, and benzbromarone, which inhibits URAT1 and reabsorption of urate in kidney. However, these medications caused hypersensitivity syndrome, hepatotoxicity or other adverse effects in specific patient populations (*6*). Therefore, it is necessary to further nominate disease-associated genes or drug targets in order to develop medications for intolerant populations. Large-scale genome-wide association studies (GWAS) have identified over 200 loci associated with serum urate levels (*7, 8*), but mechanistic understanding of the causal variants and genes underlying these loci remains partially explored, limiting the translation of genetic result to alternative medications.

For complex diseases, the presence of multiple independent associations at disease-associated locus is commonly observed (*9–12*). For example, four regions at *SLC2A9* locus have significant causal effects on serum urate levels (*13*). However, due to limitation of computational burden, the most commonly used methods for identifying causal variants, such as lead SNP extension by linkage disequilibrium (LD), genetic and epigenetic fine mapping, or colocalization (*14–17*), assume exactly one causal variant per locus (*18, 19*). As all the independent disease associations could contribute to gene regulation and trait association at the locus, these above methods may not fully map causal variants and leave a large number of loci mechanistically unresolved. Recent studies have performed the colocalization analysis of urate-associated loci with kidney expression quantitative trait loci (eQTLs), but the result could only characterize risk for tens loci and did not find the well-known targets in kidney including *SLC22A12* (encoding URAT1) and *SLC2A9* (encoding GLUT9) (*7, 20*). Therefore, it is urgent to find an effective way to overcome the issue of multiple independent associations in complex diseases and comprehensively understand the mechanism of genetic contribution.

The multiple independent associations in complex diseases also accompany the complexity of gene regulation at disease-associated locus, and hinder gene prioritization. Previous studies have shown that drug targets with genetic supports are more likely to be therapeutically valid (*21, 22*), but drug target discovery in complex diseases remains challenging. The heritability of complex diseases could be explained by the cumulative effects of many variants (*23*), which are mainly located in non-coding regions and regulate gene expression in a cell-type-specific manner (*24, 25*). Thus, cumulative-regulation-based methods to prioritize the candidate genes is expected to nominate the most probable causal genes and potential drug targets. Recent regulation-based methods have leveraged the causal gene nomination, but they are not developed for complex diseases and have certain limitations. ABC-Max only assigned one variant-gene pair with the strongest regulatory score (*26*), but overlooked the complex regulation between variants and genes (*24*) . Open Targets Genetics and H-MAGMA took into account all the variant-gene connectivity, but the connectivity is considered qualitatively rather than quantitatively (*27, 28*). The incompleteness of cumulative regulation maps, together with incomplete variant coverage and the limited cell type resolution, hinder the prioritization of causal genes. Thus, a method that systematically and accurately considers the cumulative regulation is required for prioritizing genes for complex disease.

In the current study, we propose a novel approach with locus-specific stratification and prioritization to comprehensively explore genetic contribution in complex diseases and nominate reliable disease-associated genes. Locus-specific stratification (LSS) efficiently extract all high-risk variants in addition to lead SNP based on a locus-specific background and overcome the challenge of multiple-independent-associations in complex diseases. To prioritize genes, we develop gene regulatory prioritization score (GRPS) to evaluate cumulative regulation on genes by integration high-risk variants from LSS with scATAC-seq and scRNA-seq datasets. Using this strategy, we efficiently resolved genetic risk mechanism for 118 out of 267 loci, and nominated more reliable gene targets for hyperuricemia. We further validated the regulatory potential of resolved high-risk variant on candidate causal gene by CRISPR activation (CRISPRa), and evaluated the effect of top prioritized gene on cellular urate level. Our study established a computational efficient method for GWAS interpretation of complex disease, extended genetic mechanistic understanding of hyperuricemia, and ultimately prioritizing genes with high confidence.

## Results

### Locus-specific stratification leverages interpretability of loci with multiple-independent-associations

The GWAS of serum urate level exhibited overwhelming multiple-independent-associations challenge. For the available GWAS results of serum urate level, independent association analysis revealed 22 to 128 loci with multiple independent associations, ranging from 69.95%-95.65% of all the loci (Fig. 1A). For instance, at locus chr4:9,743,616-10,243,616 with lead SNP rs3775947, in addition to lead SNP and SNPs within its high LD neighborhood, there were also numerous variants showing extremely strong trait association with -log10(p-value) even over 1000 (Fig. 1B). This suggested that such a locus may not be fully explained by only one association. Notably, at such highly associated locus, the locus-specific background is much higher than genome-wide background, suggesting a one-size-fits-all cutoff might not be appropriate.

**Fig. 1.**
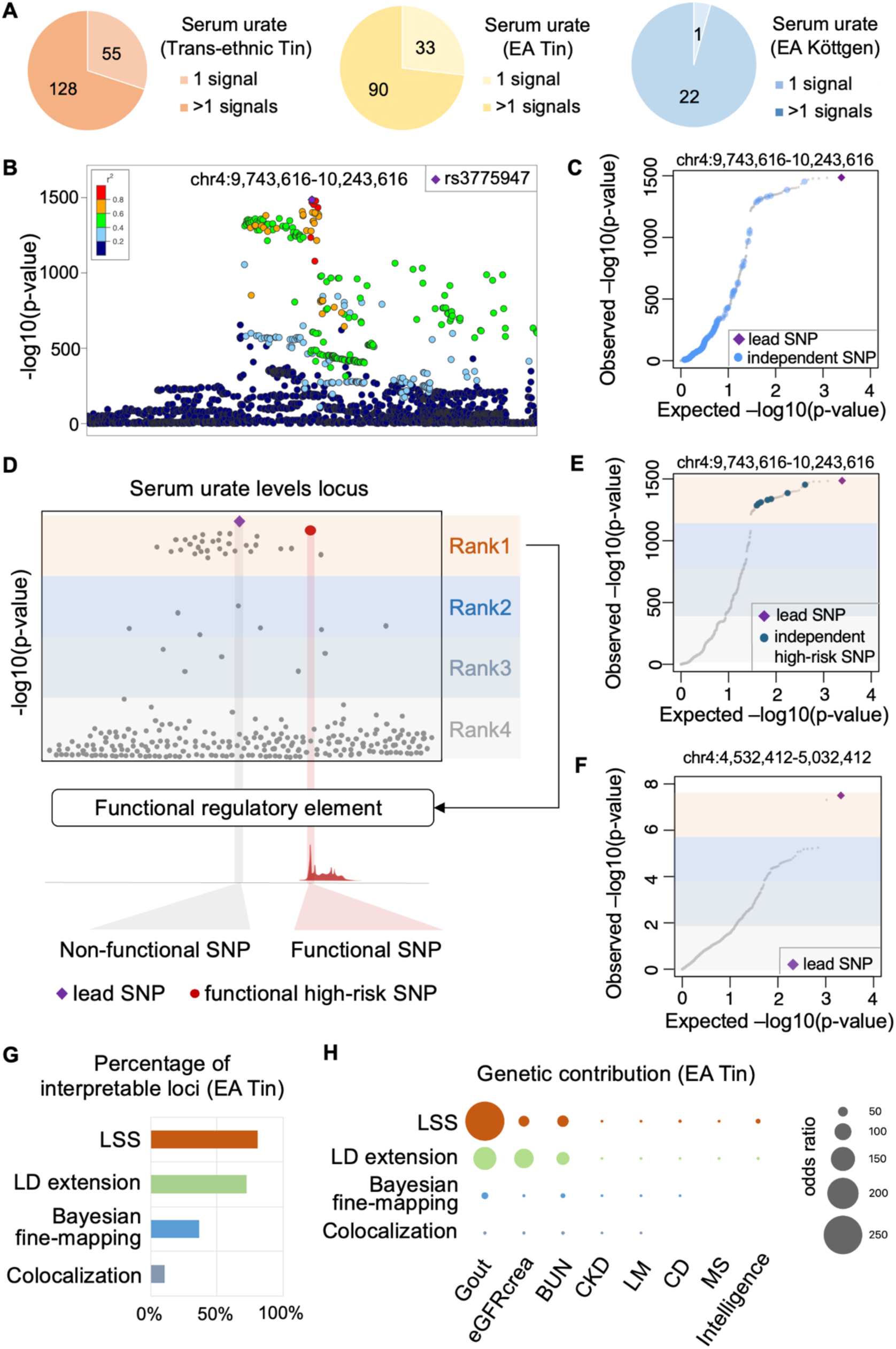
Locus-specific stratification leverages interpretability of loci with multiple-independent-associations. (**A**) Number of loci with single or multiple independent associations in three serum urate GWAS studies, respectively. (**B** and **C**) LocusZoom plot (**B**) and quantile-quantile plot (**C**) for GWAS result at locus chr4:9,743,616-10,243,616 with lead SNP rs3775947. (**D**) Overview of the LSS strategy. The variants were ranked by descending significance of variant-trait associations within each locus, and candidate high-risk variants defined as those in the top quartile. High-risk variants were further integrated with kidney scATAC-seq data to identify functional high-risk variants. (**E** and **F**) Quantile-quantile plots for GWAS result at locus with multiple independent associations (**E**) or single association (**F**). The lead SNP and independent (LD R^2^<0.8) high-risk (Rank1) SNPs were highlighted. (**G**) Percentage of interpretable loci for GWAS of serum urate (EA Tin). Four strategies including LSS, LD extension of lead SNP, Bayesian fine-mapping and kidney colocalization were used. (**H**) Odds ratios of genetic contribution for functional high-risk variants identified by four strategies. eGFRcrea, creatinine-based estimated glomerular filtration rate; BUN, blood urea nitrogen; CKD, chronic kidney disease; LM, leptin measurement; CD, Crohn’s disease; MS, multiple sclerosis.

We observed that the distribution of significancy at locus with multiple-associations tended to be reversed “L” shape with a plateau. The plateau was consisted of numerous high-risk associations, and generally harbored the top quantile associations (Fig. 1C and Fig. S1). In order to efficiently and comprehensively explore the genetic contribution, we proposed the LSS strategy to extract all high-risk associations based on locus-specific background. We ranked the variants by descending significance of variant-trait associations within each locus and took the top quartile associations as rank 1 for further analysis (Fig. 1D). LSS captured the multiple independent associations (Fig. 1E and Fig. S1) without bringing in noise at locus with single association (Fig. 1F and Fig. S2).

Previous studies highlighted that disease-associated variants enriched at tissue and cell type specific regulatory regions (*29–31*). We thus identified the target tissue and generated tissue specific single cell EpiMap to pinpoint the potential causal variants of these high-risk associations. By LD regression, we found the regulatory contribution from kidney tissue is highest (Fig. S3). We then focused on the regulatory mechanism in kidney. We generated cell type EpiMap with our previously generated kidney scATAC-seq data and integrated it with the rank 1 associations (Fig. 1D). The results showed that our strategy could explain genomic function for 220 out of 267 loci from the three studies (Table S1). When compared with other methods including high LD extension, Bayesian fine-mapping and colocalization, we found LSS could improve the interpretability of GWAS loci by 8.13% to 69.92% (Fig. 1G). Furthermore, we examined the contribution of these functional high-risk variants, and found that the variants identified by LSS contributed more highly and specifically to gout, the most common complication of hyperuricemia (Fig. 1H). These results suggested that LSS could pinpoint the functional high-risk variants for complex diseases more comprehensively and specifically.

### LSS identified variants revealed cell-type-specific contribution on complex disease

We further examined the cell types in which the identified high-risk variants could play a role in. The result showed that 77.86% of these high-risk variants exhibited chromatin accessibility in proximal tubular cells, and this fraction was followed by that in distal tubular cell types (Fig. 2A). The result is consistent with fact that urate homeostasis is maintained mainly through excretion by kidney tubules (*20*). We also found that 48.12% of the variants located in regions only accessible in one cell type, mostly in proximal tubular cells and then in distal tubules (Fig. 2B and Fig. S4). For example, at the locus chr8:75,316,533-75,816,533 with lead SNP rs2941484, the lead SNP and its proxy SNP by LD extension showed no accessibility in any kidney cell type (Fig. 2C). We found by LSS that high-risk variant rs2943549, which is in an independent association over 26kb away from the lead SNP, located in a region specifically accessible in proximal tubular cell (Fig. 2C). The phenome-wide association study (PheWAS) (*32*), which is a powerful approach to comprehensively evaluate associations between genetic variants and phenotypes, revealed that rs2943549 has the strongest association with urate-related phenotypes among the traits (Fig. 2D). In another locus chr4:9,743,616-10,243,616 with lead SNP rs3775947, a high-risk variant rs4447862 in addition to lead SNP showed accessibility, revealing an additional risk (Fig. 2E). Notably, rs4447862 demonstrated a remarkable association with uric acid (Fig. 2F), yet this risk variant could not be identified by extension of the lead SNP. These results suggested that high-risk variant from independent association could function as genomic regulation in a cell-type-specific manner, and contribute mechanistically to the genetic risk of hyperuricemia.

**Fig. 2.**
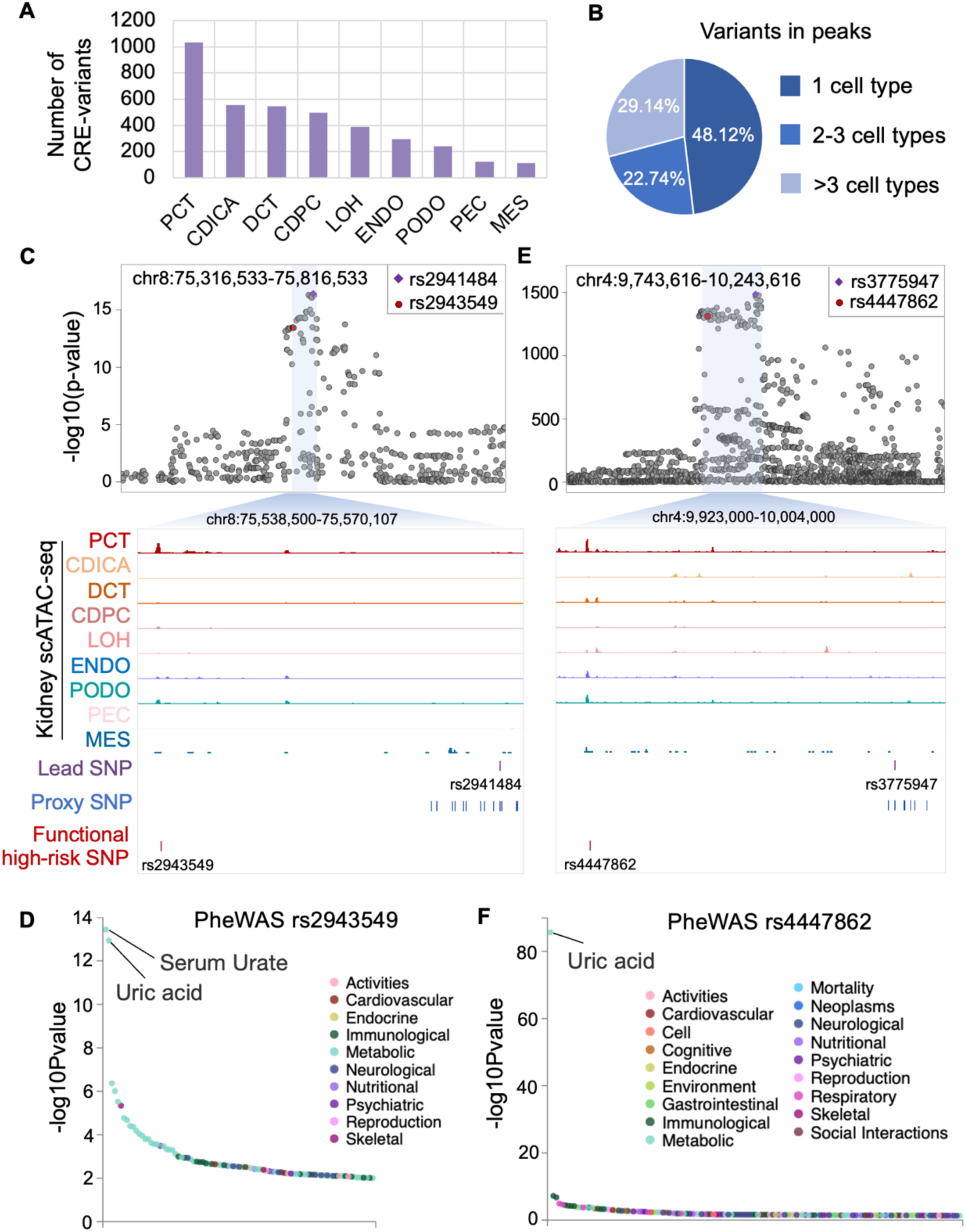
LSS reveals cell-type-dependent functional variants of hyperuricemia. (**A**) Number of high-risk variants overlapping with regulatory elements in each cell type of the kidney, including proximal convoluted tubule (PCT), collecting duct alpha intercalated cells (CDICA), distal convoluted tubule (DCT), collecting duct principal cell (CDPC), loop of Henle (LOH), endothelium (ENDO), podocyte (PODO), parietal epithelial cells (PEC), and mesangial cell (MES). (**B**) Proportion of functional high-risk variants that exert their function in unique cell types, 2-3 cell types, and multiple cell types in kidney. (**C**) LocusZoom plot of GWAS result in locus chr8:75,316,533-75,816,533 with lead SNP rs2941484 (top) and genome browser view for the highlighted region (bottom). The genome browser tracks include chromatin accessibilities in cell types from kidney scATAC-seq, the position of the lead SNP, proxy SNP (LD extension of lead SNP) and functional high-risk SNP. (**D**) The PheWAS results for rs2943549 with colors representing significantly associated diseases and traits (p<0.05). (**E**) LocusZoom plot and genome browser view for locus chr4:9,743,616-10,243,616 with lead SNP rs3775947 as (C). (**F**) The PheWAS results for rs4447862 as (D).

### Integrated regulatory network uncovers candidate causal genes of functional high-risk variants

Genetic lesions in non-coding regulatory elements contribute to disease by modulating the causal gene expression (*14*). To fully explain the regulatory mechanisms of the above functional high-risk variants, we integrated the scRNA-seq and scATAC-seq profiles of kidney to construct the transcriptional regulatory networks by identifying CREs with accessibility correlated to gene expression (‘peak-to-gene links’) (Fig. 3A and Fig. S5A). Based on the regulatory network, we identified 160 protein-coding genes linked to 232 variants in 118 loci (Table S2). Our approach interpreted mechanism for 1.4 to 5.2 fold more loci than the other approaches on the same GWAS study (Fig. 3B), and identified an additional 32 to 85 candidate genes (Fig. 3C).

**Fig. 3.**
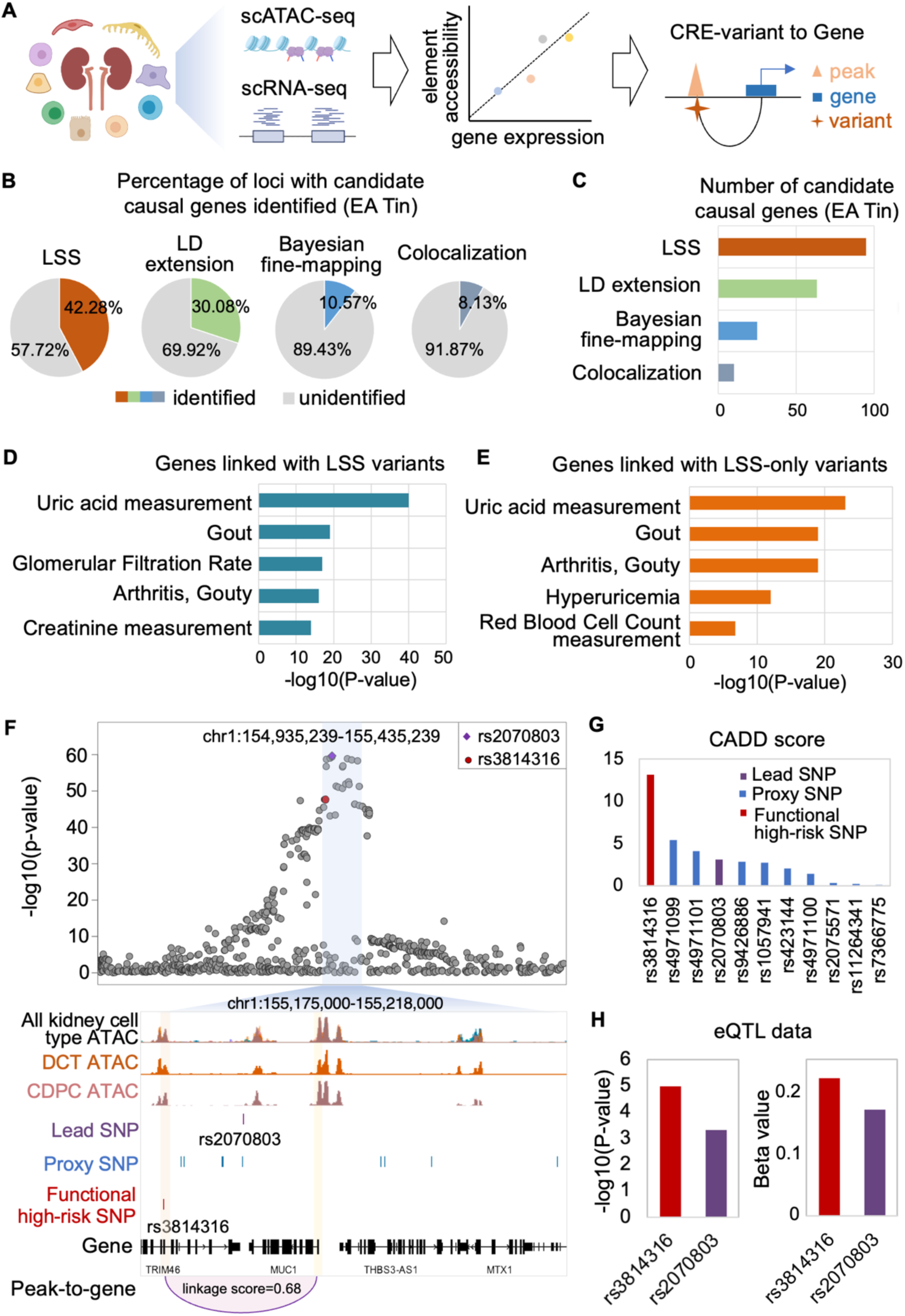
Regulatory network elucidates the regulatory mechanism for target genes and functional variants in hyperuricemia. (**A**) Overview of the construction of transcriptional regulatory networks of kidney cell types by computing the correlation between chromatin accessibility and gene expression. (**B**) Percentage of loci with kidney candidate causal genes identified in GWAS of serum urate (EA Tin). (**C**) Number of candidate causal genes identified by different strategies. (**D** and **E**) Top 5 diseases enriched for the candidate causal genes identified with all LSS high-risk variants (**D**) or LSS-only high-risk variants (**E**). (**F**) LocusZoom plot of GWAS result in locus chr1:154,935,239-155,435,239 with the lead SNP rs2070803 (top), and genome browser view of the highlighted region (bottom). The genome browser includes overlaid track of chromatin accessibilities in kidney cell types, the tracks for chromatin accessibilities in DCT or CDPC, the tracks for location of the lead SNP, proxy SNP and functional high-risk SNP, the track for gene location, and the track for the peak-to-gene linkage. rs3814316-harboring-CRE and *MUC1* promoter are marked with orange and yellow boxes, respectively. (**G**) CADD scores for lead SNP, proxy SNPs and functional high-risk SNP. (**H**) Significance and effect size of the relationship between genotypes of variants and *MUC1* expression in kidney tubule eQTL.

To examine the relevance of the identified candidate causal genes from functional high-risk variants, we performed gene function analysis. The ontology and pathway enrichment revealed that the genes were mainly involved in urate and organic anion transport (Fig. S5B). Disease enrichment analysis showed that they were involved in urate measurement and gout (Fig. 3D). Notably, genes linked with LSS-only high-risk variants were also implicated in uric acid measurement (Fig. 3E and Fig. S5C). For example, at the locus chr1:154,935,239-155,435,239, we found that independent high-risk variant rs3814316, instead of lead SNP rs2070803, functioned as a regulatory element in distal tubular cell types and linked to gene *MUC1* (Fig. 3F). *MUC1* has been shown to be important for urate level as its mutation in tubulointerstitial nephropathy patients can result in clinical manifestations of gout (*33*). In contrast, the lead SNP rs2070803 showed no regulatory capacity for kidney *MUC1* expression. In consistent with our finding, rs3814316 showed higher CADD score and cast even stronger eQTL effect on *MUC1* than rs2070803 (Fig. 3G and 3H). These results suggested that the independent high-risk variant rs3814316 identified by our study was more likely to be the causal variant for disease association at the *MUC1* locus. Our findings have not only identified candidate causal genes for disease-associated loci, but have also proposed the more plausible regulatory mechanisms for pathogenic genes. Taken together, LSS is a computational efficient method to deal with multiple-independent-association challenge of complex disease, and could better explore the mechanism of complex diseases.

### Comprehensive prioritization score nominates potential drug target genes

As there are independent high-risk variants identified by LSS, there could be complex regulation between variants and candidate causal genes. For urate-level associated loci, we observed that 42% variants could be linked to more than one gene, and 44% of the identified candidate causal genes could be regulated by more than one variant (Fig. 4A and 4B). For example, *SLC22A12*, which encodes a known urate transporter, URAT1, was regulated by up to 15 functional high-risk variants in 11 regulatory elements identified in this study at locus chr11: 64,315,390-64,815,390 with lead SNP rs71456318 (Fig. S6). In order to nominate the most probable causal genes for further targeted therapies, it is necessary to prioritize genes based on a comprehensive regulation estimation. We developed gene regulatory prioritization score (GRPS) to take full consideration of the complexity of transcriptional regulation. GRPS did not require any previous functional evidence, but rather evaluated pathogenic risk for genes objectively based on the risk level of causal variants linked to the gene, the regulation strength of causal variants on the gene, and the polygenic effects of multiple independent causal variants (Fig. 4C). Prioritizing all 160 candidate genes using GRPS showed that the top candidates were overwhelmingly supported by evidence for trait association from previous studies (Fig. 4D).

**Fig. 4.**
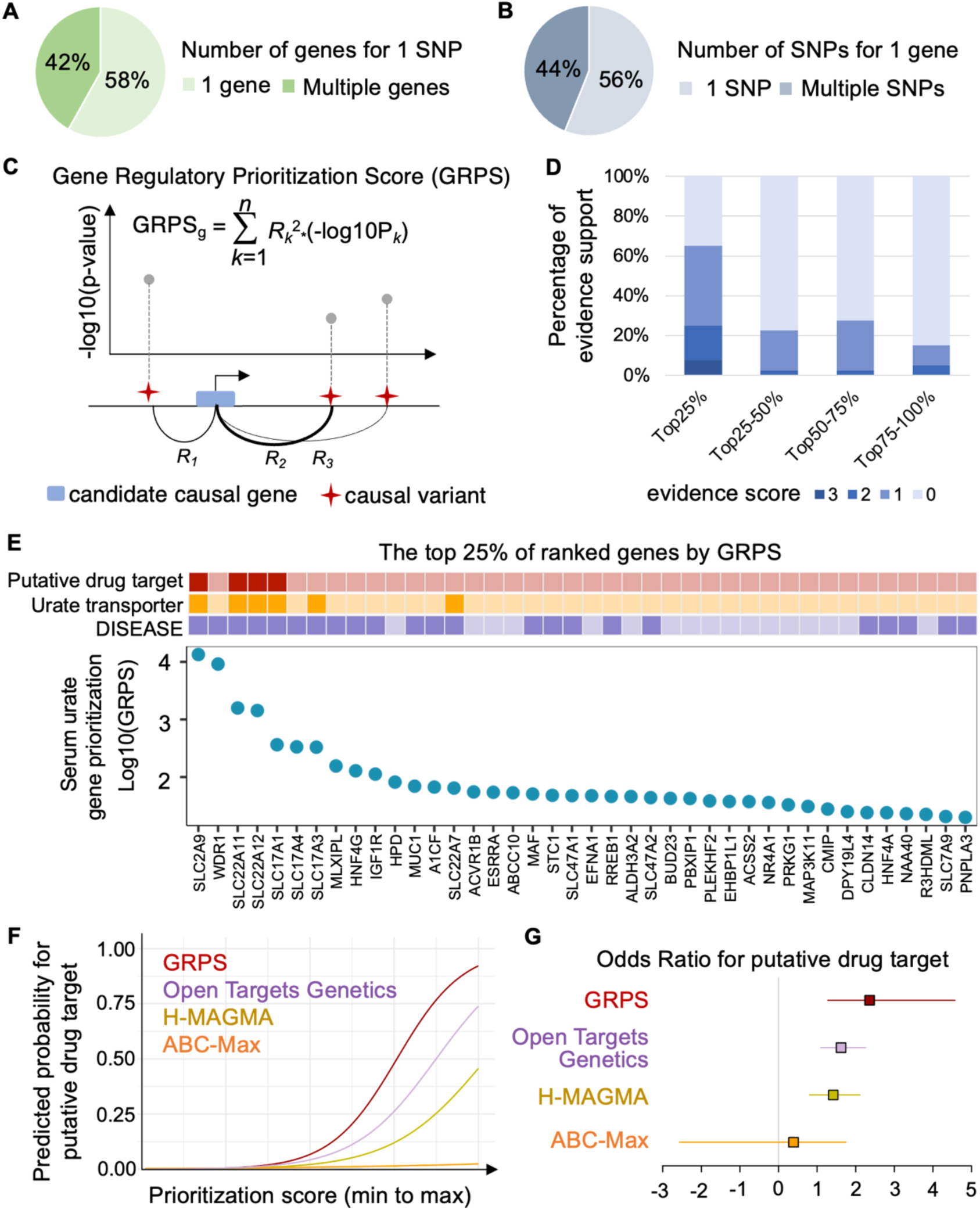
GRPS determines the priority of candidate risk genes. (**A**) Percentage of functional high-risk variants that can regulate 1 or more genes. (**B**) Percentage of gene that can be regulated by 1 or more functional high-risk variants. (**C**) Overview of GRPS strategy. For each gene, a regulatory prioritization score is calculated based on cumulative regulations (Methods). (**D**) Percentage of genes supported by prior evidence. Genes are divided into 4 groups based on GRPS from highest to lowest. (**E**) Top 25% genes ranked by the GRPS prioritization with their annotation as putative drug targets, urate transporter, and disease association by DISEASE displayed. (**F** and **G**) The performance of four different prioritizing methods in predicting the probability (**F**) and odds ratio (**G**) of putative drug targets, which was evaluated by logistic regression.

It is noteworthy that the top-ranking genes extensively enriched for known urate transporters and gene targets that are already in clinical use or clinical trials (Fig. 4E). *SLC22A11* and *SLC22A12* have been approved as confirmed drug targets for targeted therapies of hyperuricemia and gout (*34*), and *SLC2A9* and *SLC17A1* have been validated as potent targets for therapy in vivo (*35, 36*). Furthermore, the priority by GRPS ranking could more thoroughly and effectively uncover putative drug targets (Fig. 4F and 4G). For example, *SLC17A1* was ranked 5^th^ in GRPS, but was positioned at 42^nd^ in Open Targets Genetics, and was even missed out in H-MAGMA or ABC-Max (Table S3).

In addition to hyperuricemia, we also applied LSS and GRPS to other complex diseases and traits including creatinine-based estimated glomerular filtration rate (eGFRcrea), blood urea nitrogen (BUN), urinary albumin-to-creatinine ratio (UACR), gout and chronic kidney disease (CKD). The results showed that LSS generally increased the interpretability of GWAS for complex diseases and traits with multiple independent associations (Fig. S7A-D), and GRPS uncovered candidate causal genes with high credibility (Fig. S7E). All these results indicated that the approach was generally applicable to GWAS of complex diseases and could leverage the investigation of mechanism and drug target of complex diseases.

### *SLC17A4* as top nominator promoted the cellular urate transport

Among the top nominated gene targets, *SLC17A4* was the one that has not been demonstrated to be involved in urate levels (Fig. 4E). Although it was supposed that *SLC17A4* encodes an organic anion transporter, its role in cellular urate transport and regulatory mechanism have never been determined. At the GWAS locus chr6:25,559,488-26,059,488 where *SLC17A4* located, the lead SNP rs1359232 did not show regulatory potential, but the additional high-risk variant rs1165183 located at a regulatory region and linked with the 81 kb upstream gene *SLC17A4* (Fig. 5A). rs1165183-harboring-CRE was proximal tubular-specific accessible and *SLC17A4* specifically expressed in proximal tubular cells (Fig. 5A and 5B). In order to verify the regulatory potential of rs1165183-harboring-CRE on *SLC17A4*, we used CRISPRa system (*37*) to activate the activity of the regulatory element (Fig. S8A). Upon activation, we found that the expression of *SLC17A4* was significantly up-regulated (Fig. 5C).

**Fig. 5.**
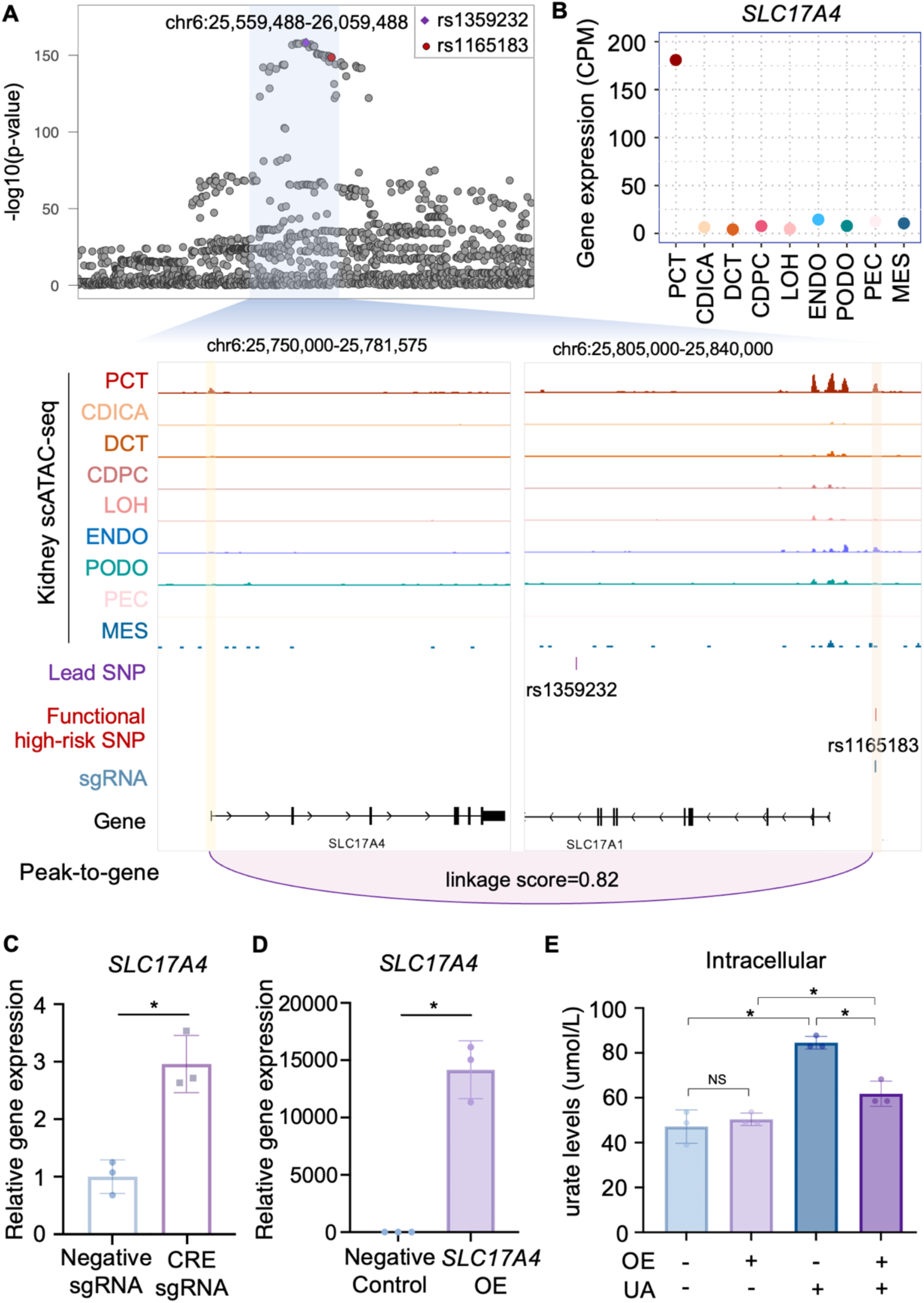
*SLC17A4* is regulated by high-risk variant and promotes the transport of uric acid. (**A**) LocusZoom plot of GWAS result in locus chr6:25,559,488-26,059,488 with lead SNP rs1359232 (top), and genome browser view of the highlighted region (bottom). The genome browser includes tracks for chromatin accessibilities in kidney cell types, the position of the lead SNP and functional high-risk SNP, the location of sgRNA designed for CRISPRa experiment, the location of genes, and the peak-to-gene linkage. rs1165183-harboring-CRE and *SLC17A4* promoter are marked with orange and yellow boxes, respectively. (**B**) Gene expression of *SLC17A4* in kidney cell types. (**C**) *SLC17A4* expression determined by qPCR in CRISPRa HEK293T lines treated with non-targeting negative sgRNA or rs1165183-harboring-CRE sgRNA (CRE sgRNA) (n=3, two-tailed Student’s t test, P-value for cells treated with negative sgRNA vs CRE sgRNA is 0.005, * indicates P-value <0.05). (**D**) *SLC17A4* expression in HEK293T cells treated with negative control plasmid and *SLC17A4* overexpression (OE) plasmid (n=3, two-tailed Student’s t test, P-value for cells treated with negative control plasmid vs *SLC17A4* OE plasmid is 0.0006, * indicates P-value <0.05). (**E**) Effects of *SLC17A4* overexpression (OE) on the intracellular urate levels of HEK293T cells (n=3, two-tailed Student’s t test, P-value for negative control cells which are untreated or treated with uric acid (UA) is 0.0012; P-value for negative control vs *SLC17A4* OE cells which are all treated with UA is 0.0033, P-value for negative control cells vs *SLC17A4* OE cells which are all untreated with UA is 0.5185, P-value for *SLC17A4* OE cells which are untreated or treated with UA is 0.0351; * indicates P-value <0.05, NS indicates not significant).

As rs1165183 was associated with urate level with -log10(p-value) as high as 148, *SLC17A4* was a strong candidate gene to affect urate level. To further determine the function of SLC17A4 on urate level, we carried out functional experimental using a sodium urate-induced model. We overexpressed SLC17A4 in HEK293T cells which did not express SLC17A4 endogenously (Fig. 5D). We then added sodium urate and monitored changes in urate levels. The results showed that overexpression of SLC17A4 significantly changed cellular urate levels upon sodium urate overload (Fig. 5E and Fig. S8B). The results confirmed that rs1165183-element could modulate the expression of SLC17A4 which affected the transport of cellular urate.

## Discussion

GWAS studies of serum urate levels have identified hundreds of genetic loci associated with hyperuricemia. However, multiple independent association in complex diseases complicates the deciphering of the mechanisms of these loci. Here, we proposed an integrated strategy that combined LSS and GRPS to deal with the multiple independent associations of complex diseases. LSS improved the functional interpretability of GWAS for complex diseases. Based on the candidate causal variants and genes revealed by LSS, GRPS considered the complexity of transcriptional regulatory networks for gene prioritization, and comprehensively and accurately nominated causal genes. Our work further investigated and confirmed the regulatory mechanism and function of *SLC17A4* locus and contributed to a theoretical foundation for the development of alternative medication in the future.

Our research has introduced a strategy for full understanding of genetic contribution in complex diseases. LSS differs from previous approaches which assume single association, it can efficiently and comprehensively extract high-risk variants from all independent associations without increasing computational burden. It significantly enhances the interpretability of GWAS results of complex diseases. For example, LSS facilitates identification of the causal genes *SLC22A12* and *SLC22A11*. In contrast, the previous colocalization analysis of serum urate GWAS and kidney eQTL did not show the two genes as causal genes (*7, 34*). *SLC22A12* and *SLC22A11* are in the locus which is highly associated with urate levels with -log10(p-value) as high as 245. URAT1 encoded by *SLC22A12* is the target of benzbromarone which is an well-known uricosuric drug that functions by increasing urate excretion in the kidney’s proximal tubule through inhibition of the dominant apical urate exchanger in the human proximal tubule (*38*). OAT4 encoded by *SLC22A11* is the target of probenecid which is approved to treat gout (*34*). All these evidences indicated *SLC22A12* and *SLC22A11* are truly causal genes for genetic association of urate level. Additionally, our study could reveal the regulatory mechanism for pathogenic genes. We identified the potential causal variant rs38814316 instead of the lead SNP for the regulation of *MUC1*, revealing an unknown regulatory mode for known pathogenic genes. Thus, LSS can be used to leverage the interpretation of GWAS of complex diseases and fully uncover the underlying mechanisms.

In evaluating the pathogenicity of all candidate causal genes, the GRPS considers all essential factors for the regulatory prioritization. This strategy reliably identifies the causal genes for complex diseases based on their fully linked genetic regulation, without the necessity of prior functional evidence. This approach is valuable for uncovering the genetic underpinnings of diseases, especially when experimental data for biological function may be limited or unavailable. Compared to other methods such as H-MAGMA, ABC-Max and Open Targets Genetics, which do not sufficiently consider the complexity of regulation at loci in complex diseases, GRPS-prioritized top genes exhibited a strong association with putative drug targets. We successfully identified 4 drug targets in kidney in the top rank. In addition to already known drug targets and our newly proposed drug target, it is interesting that *IGF1R* is among the top prioritized genes. *IGF1R* have been already approved as drug targets for treating other diseases (*34*). Our finding provides a favorable condition for drug repurposing. Overall, it is indicated that comprehensive integration of regulatory information provides a more reliable evaluation of gene pathogenicity, and can help identify promising drug target genes to facilitate drug development for treating complex diseases.

However, a potential limitation of this study is that it focused exclusively on kidney tissue that shows the highest risk for hyperuricemia. In addition to kidney tissues, we did not explore other tissues involved in urate metabolism, such as the intestine and liver, which may also contribute to the pathogenesis of hyperuricemia (Fig. S3). Subsequent applications of our proposed strategy in other tissues may expand the understanding of the pathogenesis of hyperuricemia. Importantly, this work takes an important step forward by providing comprehensive functional annotation of risk loci in complex diseases. We have also tested our strategy to the GWAS of other kidney complex diseases or traits and demonstrated the robust applicability of our strategy and its power in tackling the complexities of genetic associations. With the ongoing expansion of GWAS interpretation, the effective integration of LSS and GRPS will offer further opportunities and possibilities for disease prediction, treatment and prevention.

## Data Availability

The kidney scATAC-seq publicly available data used in this study are available in the Gene Expression Omnibus (GEO) under GSE166547. The kidney scRNA-seq data used in this study are publicly available in the GEO under GSE131882. Kidney tubule eQTL derives from NephQTL browser (https://www.nephqtl.org). Epigenomic profiles from EpiMap is available at http://compbio.mit.edu/epimap/. GWAS datasets used in this study are downloaded from CKDGen Consortium (https://ckdgen.imbi.uni-freiburg.de).

## Funding

J.Y. is grateful to support from The Open Project of Jiangsu Provincial Science and Technology Resources (Clinical Resources) Coordination Service Platform JSRB2021-01. S. S. is grateful to support from National Natural Science Foundation of China (82170723).

## Author Contributions

J.Y. conceived of and designed the project. J.Z., Y.G., and L.G. performed research. J.Z., L.X., Q.L., K.W., and Q.W. contributed to data analysis. J.Y., J.Z., S.S., and Z.Q. wrote the manuscript with input from all authors. All authors read and approved the final submission of the manuscript.

## Conflict of interests

The authors declare that they have no competing interests.

## Data and materials availability

The kidney scATAC-seq publicly available data used in this study are available in the Gene Expression Omnibus (GEO) under GSE166547 (*39*). The kidney scRNA-seq data used in this study are publicly available in the GEO under GSE131882 (*40*). Kidney tubule eQTL derives from NephQTL browser (https://www.nephqtl.org). Epigenomic profiles from EpiMap is available at http://compbio.mit.edu/epimap/. Main code used in processing and analysis of the data in this article is available at https://github.com/NJU-labyang/hyperuricemia. Any code not provided there will be made available upon request.

## Notes

### Competing Interest Statement

The authors have declared no competing interest.

