## Supplementary Materials for "Locus-specific stratification and prioritization unveil high risk genes underlying hyperuricemia"

**The PDF file includes:**

Materials and Methods
Figs. S1 to S8
References

### Materials and Methods

#### GWAS data collection and multiple independent associations analysis

GWAS datasets of serum urate, eGFRcrea, BUN, UACR, gout and CKD used in this study are downloaded from CKDGen Consortium (https://ckdgen.imbi.uni-freiburg.de). GWAS datasets of leptin measurement, crohn disease, multiple sclerosis and intelligence are downloaded from GWAS Catalog (http://ftp.ebi.ac.uk/pub/databases/gwas/summary_statistics/). To obtain the independent associations of GWAS loci, we used FUMA (*1*) (https://fuma.ctglab.nl/snp2gene) with default parameters and $r^2$ threshold to define independent significant SNPs is < 0.8. LocusZoom plots of the interest regions were visualized with LocusZoom (http://locuszoom.org).

#### LDSC regression analysis

We downloaded chromatin segmentation of different tissues from EpiMap (*2*), and active enhancer regions were used as functional categories for LDSC (*3*). LDSC regression analysis was performed with LDSC version 1.0.1 by following the tutorial (https://github.com/bulik/ldsc/wiki). The results of LDSC regression coefficient Z-scores were visualized with the R package pheatmap version 1.0.12.

#### Locus-specific stratification strategy and functional variants identification

Previous studies reported a total of 267 unique lead SNPs in three GWASs on serum urate level (*4, 5*). For each locus within 500kb of the reported lead SNP, we quartered the significance of variant-trait associations from highest to lowest. Variants with the top quartile associations were extracted as rank 1 high-risk variants and were used for subsequent analysis. Open chromatin regions of kidney cell types obtained from our kidney scATAC-seq data for functional annotation. Functional SNPs were identified by overlapping high-risk variants itself or its LD extension with regulatory elements of kidney cell types. For those high-risk variants that did not overlap with regulatory elements by itself, we performed LD extension of the variants. Proxy SNPs were extracted with $r^2 > 0.8$ using LDlinkR version 1.1.2 (*6*). A total of 1328 functional high-risk variants were identified.

#### Other fine-mapping methods

For LD extension, proxy SNPs were determined as SNPs within high LD ($r^2 > 0.8$) with lead SNPs. Bayesian fine-mapping was performed as described previously (*7*). Specially, for each locus, we first pre-filtered variants in at least low LD ($r^2 > 0.1$) with the lead SNPs. We calculated approximate Bayes factors (aBF) for each variant using the effect estimates (β) and standard errors (SE), assuming prior variance w=0.04. We calculated the posterior probability of association (PPA) by dividing the aBF for each variant by the sum of aBFs for all variants included in the signal. We then defined the 99% credible set as the smallest set of variants that added up to 99% PPA. The SNPs extracted by LD extension and Bayesian fine-mapping were then annotated using open chromatin regions of kidney cell types. Functionally annotated variants were categorized as functional high-risk variants in these two methods. Colocalization SNPs of urate-associated loci with kidney eQTLs were obtained from published manuscript and considered as the functional high-risk variants in colocalization (*4*).

**Comparation of other fine-mapping methods to LSS**

GWAS results of serum urate (EA Tin) were used to compare all other fine-mapping methods to LSS. Interpretable loci are defined as loci in which functional high-risk variants can be identified. For enrichment analysis of genetic contribution for functional high-risk variants, a random equal number of non-significant variants in the serum urate GWAS summary were used as control. The odds ratio was calculated as the enrichment of disease association from the functional high-risk variants compared to that of random control variants.

**Annotation of the variants**

PheWAS plot is generated from https://atlas.ctglab.nl/PheWAS. CADD score is obtained from https://cadd.gs.washington.edu.

**scATAC-seq and scRNA-seq analysis**

For the pre-processing of the 10x-based kidney scATAC-seq data, reads were aligned to the hg38 human genome by using the "cellranger-atac count" function in cellranger-atac version 2.0.0. Quality control measurements and filtering were conducted using the ArchR version 1.0.2 (*8*) with default parameters. We re-analyzed the kidney scRNA-seq dataset by using scRNA-seq pipeline in Seurat version 3.0.0 (*9*) and annotating with markers reported in previous manuscript (*10*).

**Construction of transcriptional regulatory network**

We leveraged the integrated kidney scRNA-seq and scATAC-seq data by ArchR version 1.0.2 to compute the linkage score between chromatin accessible regions and gene expression (‘peak-to-gene links') by using the "getPeak2GeneLinks" function in ArchR with default parameters. We further filtered out the linkages of peak-to-gene with high linkage score with abs (linkage score) > 0.6 and with genes expressed in not less than 5% cells.

**Gene regulatory prioritization score**

In order to prioritize the risk genes, we calculated a GRPS for each candidate gene by considering cumulative regulatory factors including the risk significancy of its linked causal variants, the regulation strength from its linked causal variants, and the polygenic effects of multiple independent variants as follow:

$$\mathrm{GRPS}_g = \sum_{k=1}^{n} R_k{}^2 * (-\log 10 \mathrm{P}_k)$$

where $\mathrm{GRPS}_g$ is the cumulative regulatory prioritization score of gene *g*, *n* is the number of linked risk-peaks (peaks with high-risk variants identified by LSS) for gene g; *R* is the peak-to-gene linkage score for peak *k* on gene *g*; P is the highest disease association of high-risk variant in peak *k.* GRPS value were used to prioritize 160 candidate genes identified by LSS. The top 25% of ranked genes by GRPS prioritization were visualized with the ggplot2 version 3.1.1.

**Other gene prioritization methods**

For H-MAGMA (*11*), exonic and promoter SNPs were assigned to the genes in which they reside, while intronic and intergenic SNPs were coupled to their regulated genes based on high linkage score detected in peak-to-gene links. Then we ran H-MAGMA version 1.08 with the default parameter and obtained the gene prioritization. The priority by Open Targets Genetics (*12*) is downloaded by searching for 'urate measurement' at Open Targets (https://genetics.opentargets.org). The priority by ABC-Max with keyword 'UA' (uric acid) is downloaded from Supplementary Table 10 of previous manuscript (*13*).

**Performance of gene prioritization methods in predicting potential drug targets**

Drug targets for treatment of hyperuricemia and gout were obtained from the Supplementary Table 1 of previous manuscript (*14*). To compare the performance of different gene prioritization methods in predicting potential drug targets, the priority scores obtained by each method are first subjected to quantile normalization using the R package preprocessCore version 1.60.2. Predicted probability and odds ratio for putative drug targets of priority genes by different strategies was calculated using logistic regression from "glm" in R package stats version 4.2.2. The predictor of the logistic regression model was the normalized priority score of gene, and the response of the model was whether the gene was drug target (0: no; 1: yes).

**Gene enrichment analysis**

For identified candidate causal genes, gene ontology and pathway enrichment analysis and DisGeNET enrichment analysis were performed with Metascape version 3.5 (http://metascape.org). Supporting evidence for candidate genes came from three sources: genes associated with hyperuricemia or gout in DISEASE (https://diseases.jensenlab.org/), genes were the strongest association with urate-related phenotypes in PheWAS (https://atlas.ctglab.nl/PheWAS), or genes for which mechanisms were explored in previous studies (*15-17*). The evidence score is sum score of above supports.

**CRISPR activation**

CRISPRa experiments were performed as previously described procedure (*18-20*). Lenti dCAS-VP64_Blast (Addgene, #61425) and lenti MS2-P65-HSF1_Hygro (Addgene, #61426) were packed as lentivirus and transfected to HEK293T cells to generate stable synergistic activation mediator (SAM) cell lines under 200 μg/mL Hygromycin (Thermo Fisher Scientific, 10687010) and 5 μg/mL Blasticidin (Thermo Fisher Scientific, A1113903) selection for 14 days. sgRNA of rs1165183-harboring-CRE was designed by IDTdna (https://sg.idtdna.com/site/order/designtool/) and the sgRNA of non-human-genome-targeting was derived from human GeCKOv2 library (*18*) as the negative control

(Table S4). The sgRNA were then inserted into lenti sgRNA (MS2) Puro backbone (Addgene, #73795) using BsmBI (NEB, R0739S) sites. The lentivirus of sgRNA was prepared and transfected into stable SAM cell lines under 2 μg/mL Puromycin (Thermo Fisher Scientific, A1113803) selection for 48 hours.

**Quantitative real-time Polymerase Chain Reaction (qRT-PCR)**

Total mRNA was extracted using TaKaRa MiniBEST Universal RNA Extraction Kit (Takara, 9767) following manufacturer's instruction. The RNA obtained (500 ng) was reverse-transcribed using PrimeScript™ RT Master Kit (Takara, RR036A) following the manufacturer's instruction. mRNA levels for *SLC17A4* were measured by qRT-PCR using Power SYBR™ Green PCR (Applied Biosystems™, 4367659) according to the manufacturer's instruction (Table S4).

***SLC17A4* overexpression**

The mammalian expression vector pcDNA3.1-GFP and pcDNA3.1-SLC17A4 was purchased from GenePharma. Transfection of negative control plasmid and overexpressing plasmid were performed using Lipofectamine 3000 (Invitrogen, L3000015) according to the manufacturer's instruction. For transfection, HEK293T cells were seeded in 24-well plates at ~50% confluency, and then transfected with 500 ng plasmids for each well. After 24 hours, RNA was extracted, then reverse-transcribed and *SLC17A4* gene expression was measured by qRT-PCR.

**Determination of urate levels**

The negative control and *SLC17A4*-overexpressing HEK293T cells were treated with 80 ug/mL uric acid sodium salt (Sigma, U2875). The urate content of the supernatants and broken cells was detected using the urate kit (Nanjing Jiancheng Bioengineering Institute, C012-2-1) after incubation for 24 hours in a 37°C incubator. Three biological replications were carried out for each group.

**Statistical Analysis**

The following statistical tests were performed by two-tailed Student's t test (Fig. 5C, Fig. 5D, Fig. 5E, Fig. S8B). Data on Fig. 5C, Fig. 5D, Fig. 5E, Fig. S8B are mean ± SD.

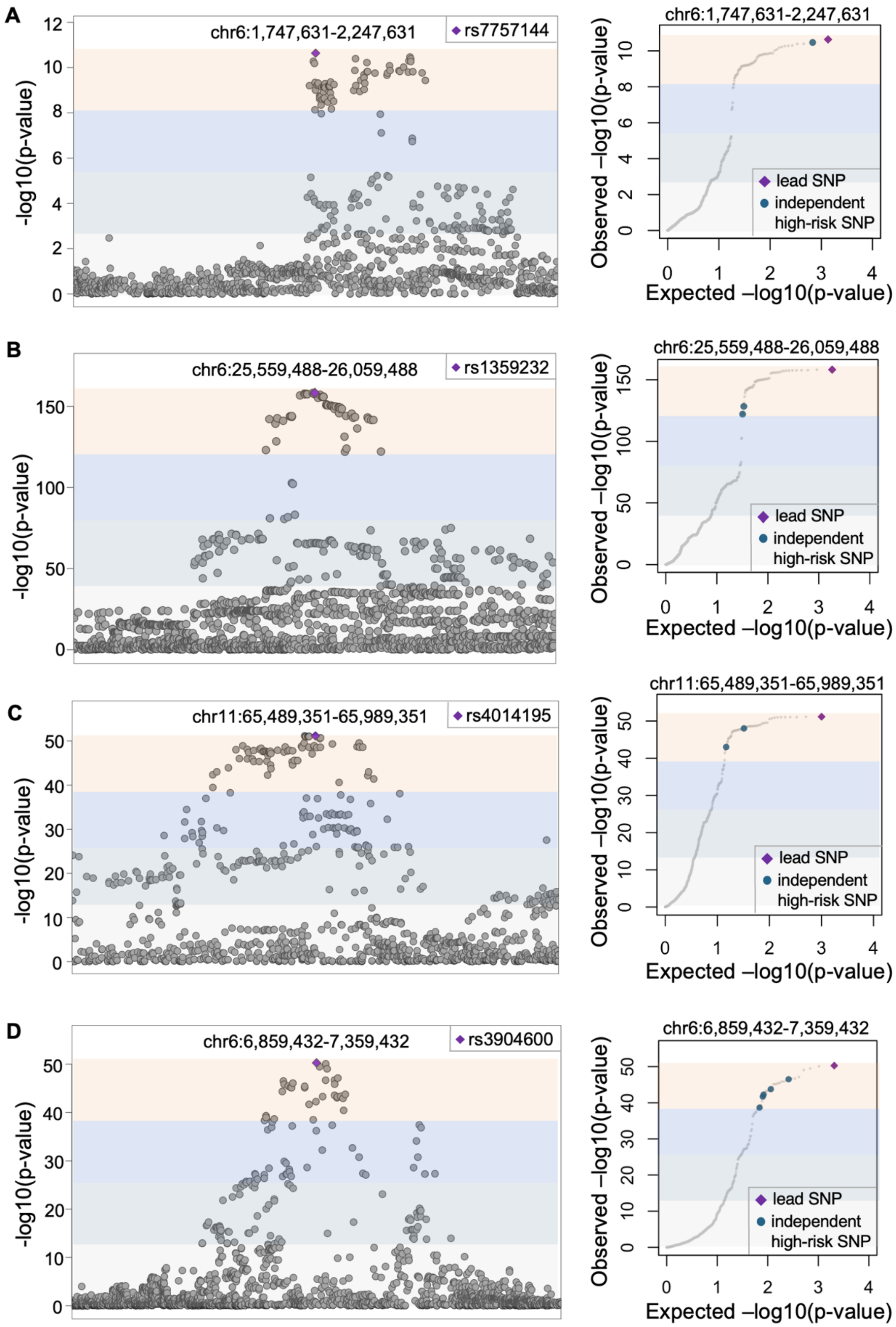
A
chr6:1,747,631-2,247,631
rs7757144
-log10(p-value)
Observed –log10(p-value)
Expected –log10(p-value)
lead SNP
independent high-risk SNP
B
chr6:25,559,488-26,059,488
rs1359232
C
chr11:65,489,351-65,989,351
rs4014195
D
chr6:6,859,432-7,359,432
rs3904600

**Fig. S1 Loci of serum urate GWAS with multiple independent associations.**

LocusZoom plots (left) and quantile-quantile plots (right) for GWAS results in loci with multiple independent associations, where chr6:1,747,631-2,247,631 with lead SNP rs7757144 (**A**), chr6:25,559,488-26,059,488 with lead SNP rs1359232 (**B**), chr11:65,489,351-65,989,351 with lead SNP rs4014195 (**C**) and chr6:6,859,432-7,359,432 with lead SNP rs3904600 (**D**), respectively.

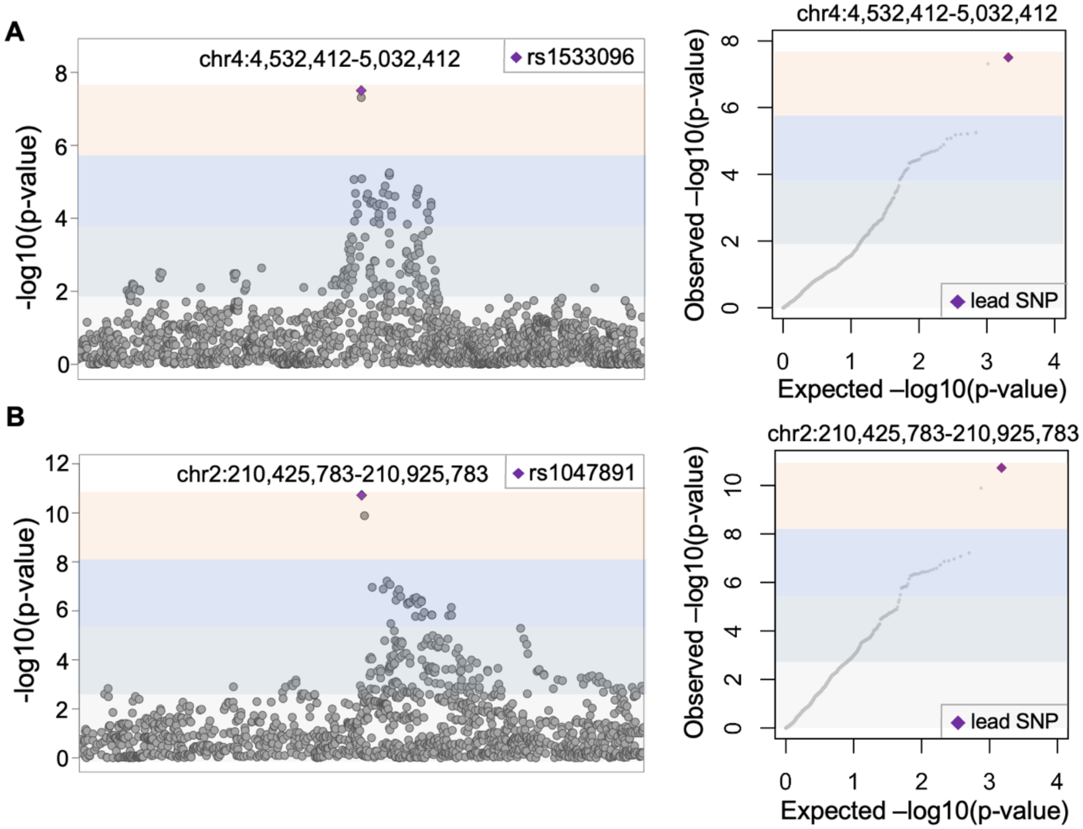


**Fig. S2 Loci of serum urate GWAS with single association.** LocusZoom plots (left) and quantile-quantile plots (right) for GWAS results at loci with single association, where chr4:4,532,412-5,032,412 with lead SNP rs1533096 (**A**), chr2:210,425,783-210,925,783 with lead SNP rs1047891 (**B**), respectively.

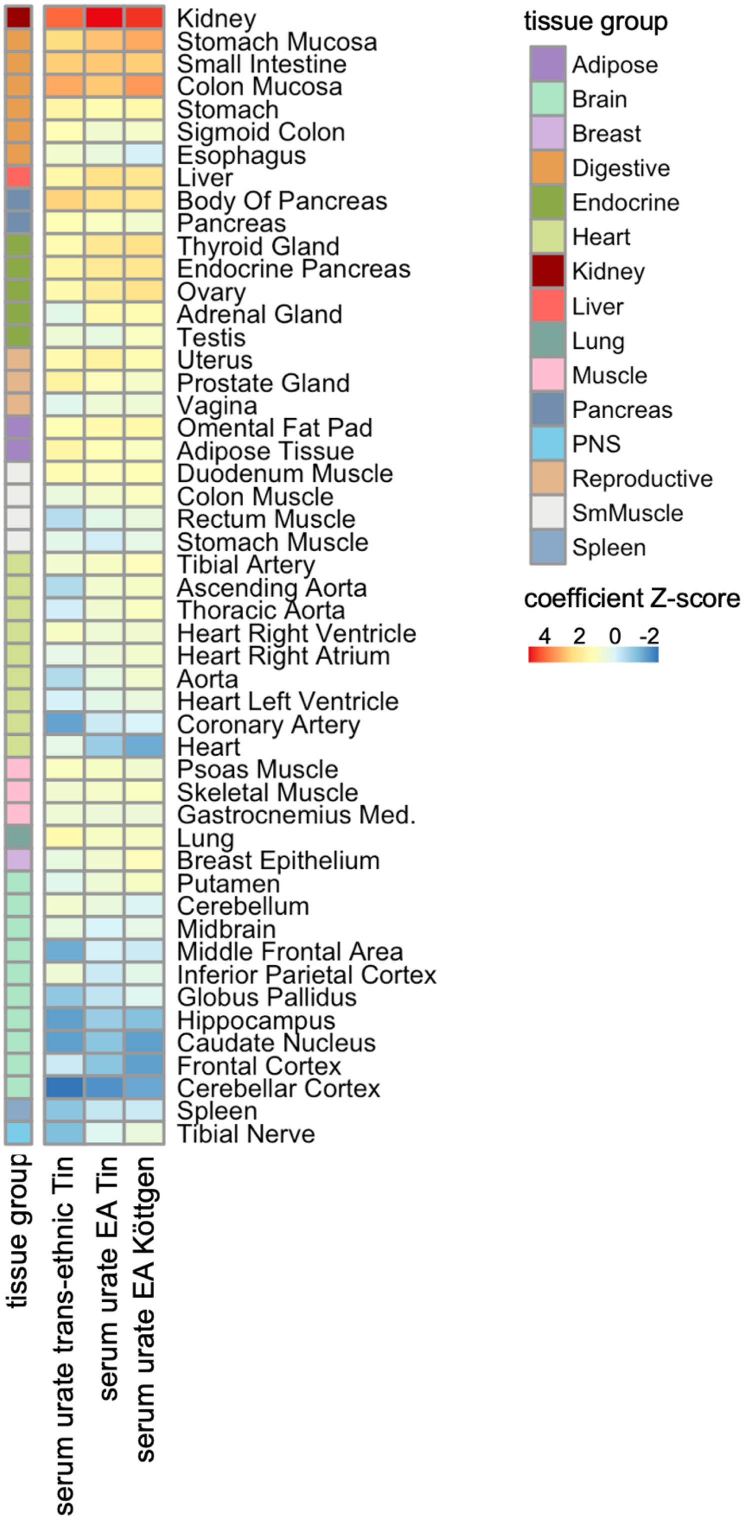


**Fig. S3 Tissue enrichment for genetic risk of serum urate.** A heatmap displaying LD regression coefficient Z-scores for human tissues across three serum urate GWAS studies analyzed. Data are annotated by groups of tissues.

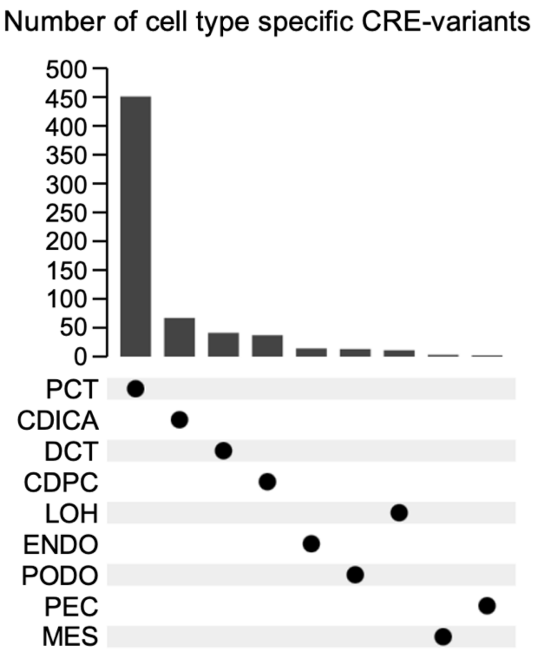


**Fig. S4 Cell type contribution for genetic risk of serum urate.** Number of high-risk variants overlapping with regulatory elements in unique kidney cell type.

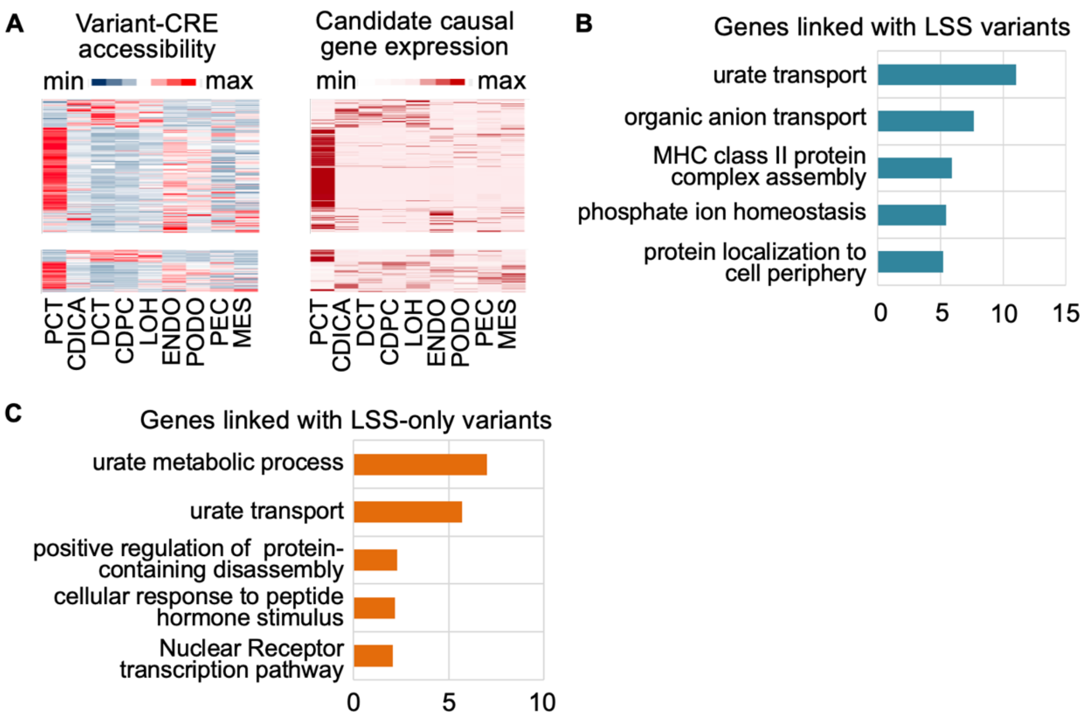


**Fig. S5 Identification of candidate causal genes for functional variants.** (**A**) Peak-to-gene links in kidney cell types including positive and negative regulation. Heatmaps showed the chromatin accessibility of regulatory elements and gene expression in peak-to-gene links, respectively. The color represents z-score. (**B** and **C**) Top 5 enriched gene ontology and pathway terms on candidate causal genes linked with LSS (**B**) or LSS-only (**C**) high-risk variants.

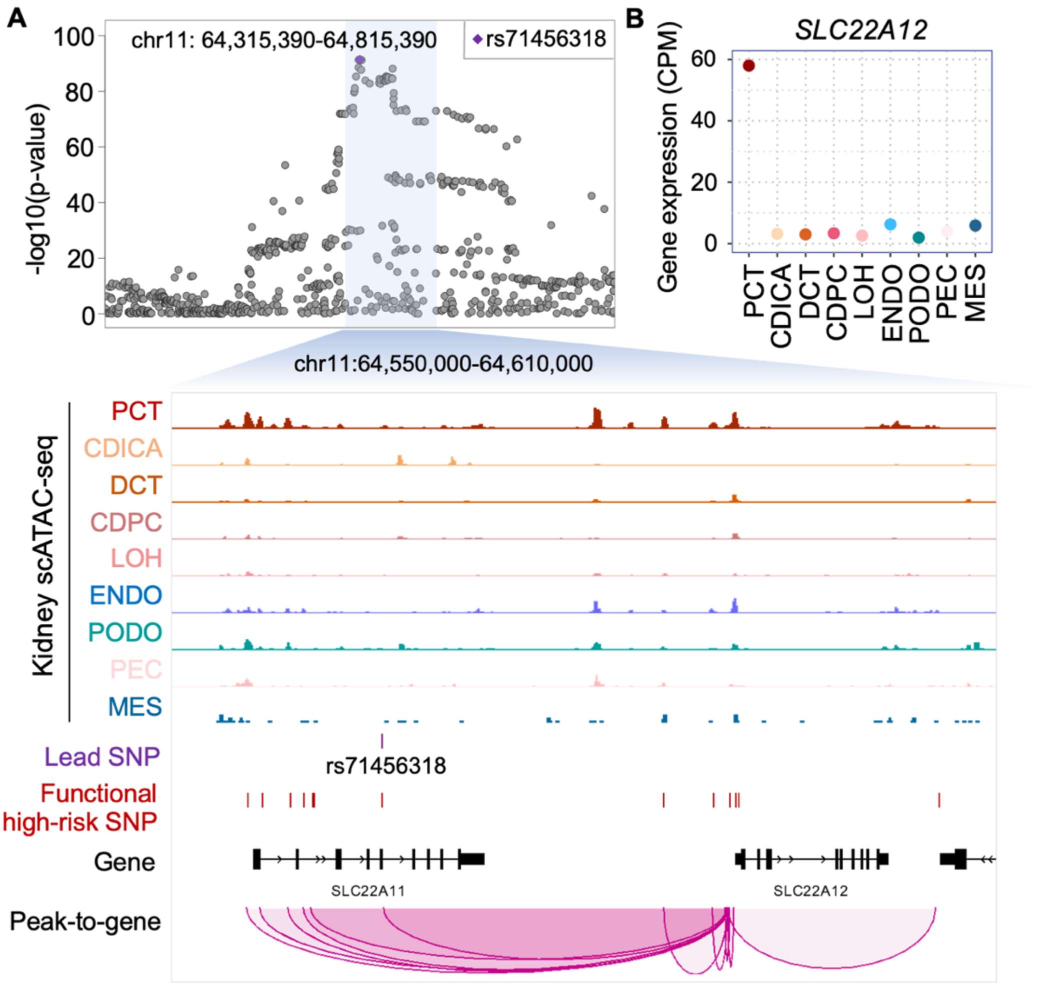


**Fig. S6 The complex regulation between variants and candidate causal genes.** (**A**) LocusZoom plot of GWAS result at locus chr11: 64,315,390-64,815,390 with lead SNP rs71456318 (top), and genome browser view of the highlighted region (bottom). The genome browser includes tracks for chromatin accessibilities in kidney cell types, the position of the lead SNP and functional high-risk SNPs, the location of genes, and the peak-to-gene linkage. (**B**) Gene expression of *SLC22A12* in kidney cell types.

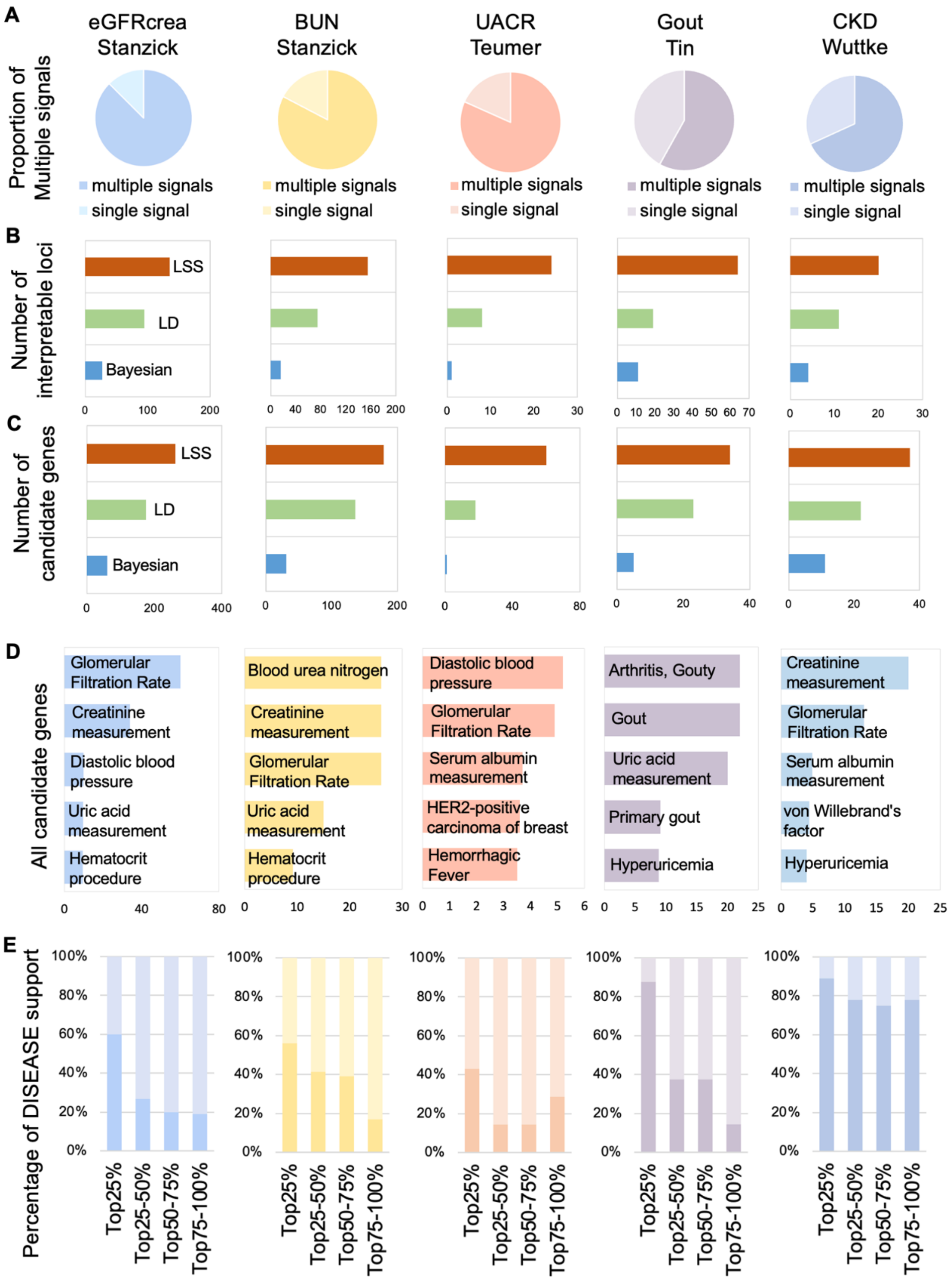


**Fig. S7 Application of LSS and GRPS to other GWAS studies.** (**A**) Proportion of loci with multiple independent associations in GWAS of eGFRcrea, BUN, UACR, gout and CKD. (**B**) Number of interpretable loci in five diseases and traits among different strategies. (**C**) Number of candidate risk genes identified by different strategies. (**D**) Top

5 enriched diseases terms on candidate causal genes regulated by functional variants. (**E**) Risk genes are divided into 4 groups based on GRPS from highest to lowest, and the bar chart shows the percentage of genes supported by disease association from DISEASE displayed.

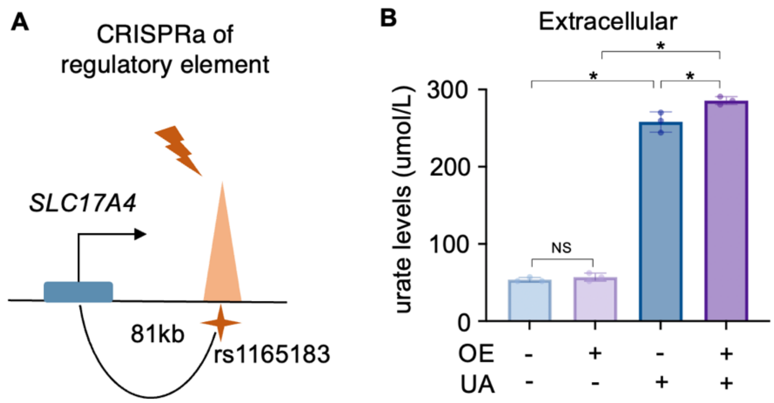


**Fig. S8 Validating the regulatory and functional mechanisms of the risk gene.** (**A**) Experimental scheme for CRISPRa with rs1165183-harboring-CRE. (**B**) Effects of *SLC17A4* overexpression on the extracellular urate levels of HEK293T cells (n=3, two-tailed Student's t test, P-value for negative control cells which are untreated or treated with uric acid (UA) is <0.0001; P-value for negative control vs *SLC17A4* OE cells which are all treated with UA is 0.0268, P-value for negative control cells vs *SLC17A4* OE cells which are all untreated with UA is 0.3738, P-value for *SLC17A4* OE cells which are untreated or treated with UA is <0.0001; * indicates P-value <0.05, NS indicates not significant).
